# Integrated single cell analysis of human lung fibrosis resolves cellular origins of predictive protein signatures in body fluids

**DOI:** 10.1101/2020.01.21.20018358

**Authors:** Christoph H. Mayr, Lukas M. Simon, Gabriela Leuschner, Meshal Ansari, Philipp E. Geyer, Ilias Angelidis, Maximilian Strunz, Pawandeep Singh, Nikolaus Kneidinger, Frank Reichenberger, Edith Silbernagel, Stephan Böhm, Heiko Adler, Anne Hilgendorff, Michael Lindner, Antje Prasse, Jürgen Behr, Matthias Mann, Oliver Eickelberg, Fabian J. Theis, Herbert B. Schiller

## Abstract

Single cell genomics enables characterization of disease specific cell states, while improvements in mass spectrometry workflows bring the clinical use of body fluid proteomics within reach. The correspondence of cell state changes in diseased organs to peripheral protein signatures is currently unknown. Here, we leverage single cell RNA-seq and proteomic analysis of large pulmonary fibrosis patient cohorts to identify disease specific changes on the cellular level and their corresponding reflection in body fluid proteomes. We discovered and validated transcriptional changes in 45 cell types across three patient cohorts that translated into distinct changes in the bronchoalveolar lavage fluid and plasma proteome. These protein signatures correlated with diagnosis, lung function, smoking and injury status. Specifically, the altered expression of a novel marker of lung health, CRTAC1, in alveolar epithelium is robustly reflected in patient plasma. Our findings have direct implications for future non-invasive prediction and monitoring of pathological cell state changes in patient organs.

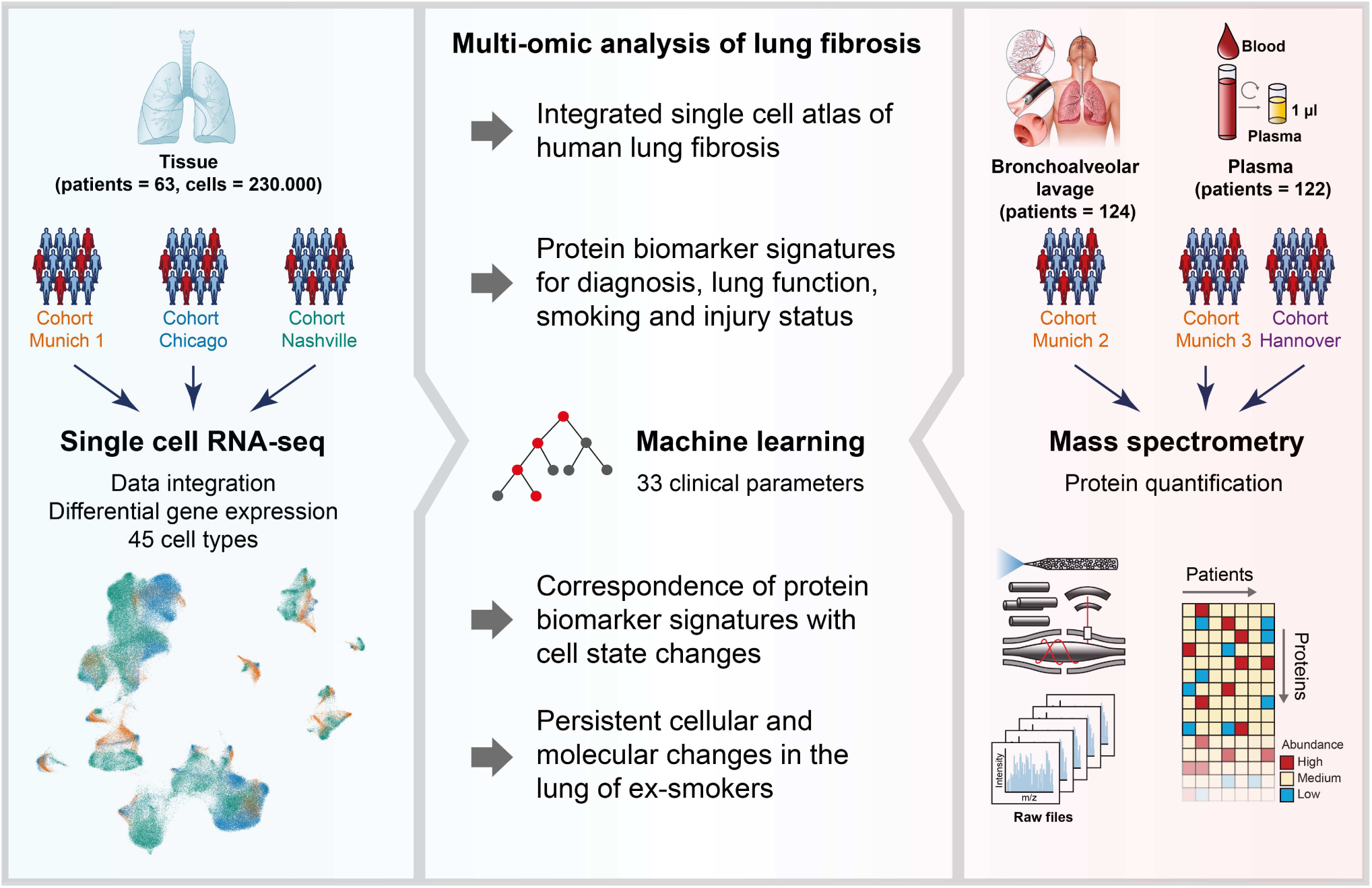

## Main

The accumulation and persistence of scar tissue in fibrotic diseases such as pulmonary fibrosis, liver cirrhosis and cardiovascular disease is among the most severe clinical issues, causing an estimated 45% of all deaths in the developed world^1^. Interstitial lung diseases (ILD) are a heterogeneous group of diseases ultimately leading to pulmonary fibrosis, destruction of the lung parenchyma and respiratory failure. Several potential risk factors have been identified, including genetic predisposition^2^, smoking^3^, infections (e.g. viruses)^4^, aging^5^, and autoimmunity^6,7^. Addressing the heterogeneity of ILD entities, disease progression and prognosis, and the currently unpredictable occurrence of acute exacerbations of disease, require new molecular approaches for personalized patient monitoring.

The recent surge of innovation in single cell genomics enables an entirely novel cell type specific viewpoint on pathological changes in disease. Based on these new technologies, the Human Cell Atlas project aims at building a comprehensive reference map of all human cells as a basis for understanding fundamental human biological processes and diagnosing, monitoring, and treating disease^8^. This includes recent international efforts towards building a human Lung Cell Atlas in health and disease^9^. A first draft of the cellular composition of mouse and human lung has been established^10–14^, and several recent single cell profiling studies reported cellular and molecular changes associated with pulmonary fibrosis^15–18^. However, this nascent draft of a human Lung Cell Atlas currently lacks extension into the complexity of the proteome layer.

As disease trajectories in ILD patients are often highly variable, protein signatures in patient body fluids promise improved personalized treatment and longitudinal monitoring of patients^19,20^. The transcriptomic and proteomic changes in endstage ILD patient lung tissue have been resolved using microarrays, RNA-sequencing and mass spectrometry^6,21,22^. Furthermore, first gene and protein expression signatures in ILD bronchoalveolar lavage (BAL), which is obtained during bronchoscopy have been analyzed^23,24^. However, it is unclear which cellular and molecular processes in the lung correspond to these biomarker signatures, representing a tissue or fluid average which does not resolve cellular composition and disease specific cell states.

In this work, we explore the idea that protein signatures found in bronchoalveolar lavage and plasma, both of which are accessible for longitudinal monitoring of patients, can be used to predict pathological cell state changes in the lung. We (1) derived cell state changes in human lung fibrosis at cellular resolution and (2) integrated our data with two independent but complementary datasets to establish robust differential gene expression analysis for all major cell types of the human lung and assess reproducibility across patient cohorts. We also used (3) state of the art mass spectrometry workflows to quantify the bronchoalveolar lavage and plasma proteome compositions in patients from several independent large-scale ILD cohorts. A (4) multi-modal analysis strategy enabled integration of cell state descriptions on single cell level with transcriptomic tissue bulk measurements, the lavage and plasma proteomes, and associated clinical meta- data. Our analysis dissects human lung fibrosis at the single cell level, defining robust differential gene expression profiles across multiple studies for ILD. Using machine learning, we show that fluid proteome signatures are predictive of specific cell state changes in the lung and discover protein biomarker signatures associated with diagnosis, lung function, and smoking and injury status.

## Results

### An integrated single cell atlas of human lung fibrosis

To analyze transcriptional changes in lung fibrosis at cellular resolution, we obtained whole lung parenchyma single cell suspensions using non-fibrotic control tissues from 11 donors and endstage lung fibrosis tissues from three ILD patients. Dimension reduction was used to visualize a data manifold representing the gene expression space of 41,888 single cells (Fig. 1a, b; control, n=11; ILD, n=3)(Fig. S1a, b). We generated subsets of the whole lung parenchyma datasets for COL1A2+ stromal cells (Fig. 1c and Fig. S1c-e), EPCAM+ epithelial cells (Fig. 1d and Fig. S1f-h), CLDN5+ endothelial cells (Fig. 1e and Fig. S1i-k), and CD45+ leukocytes (Fig. 1f and Fig. S1l-n). From these subsets we derived cluster identities (Fig. S1; Table S1) that were manually annotated using previously established single cell signatures in the human lung^10,11^. We observed some clusters that were mainly present in fibrosis and therefore termed them as ‘activated’ cell type identities (Fig. 1b, Fig. S1). The final annotation revealed 45 cell type identities, characterized by unique marker gene expression profiles (Fig. 1g-j) that were to some extent preserved in endstage fibrosis.

**Figure 1.**
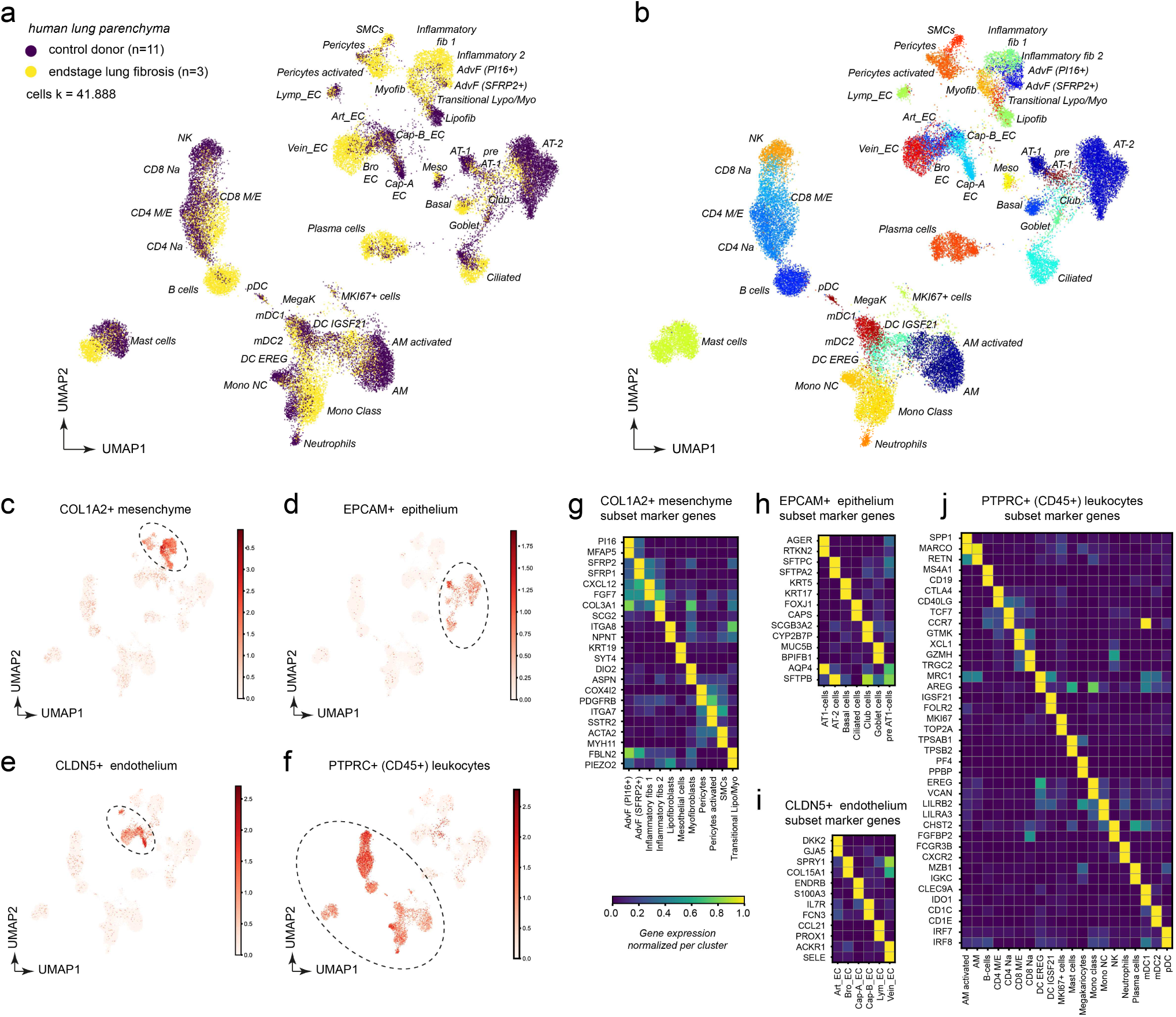
Single cell analysis of human lung parenchyma in health and disease reveals 45 distinct cell type identities and their marker genes. (a, b) Dimension reduced single cell transcriptomic data is visualized through Uniform Manifold Approximation and Projection (UMAP). The color code illustrates the disease status (a) and cell type identity (b). (c-f) The indicated marker genes were used to select clusters for subsetting into stromal cells (c), epithelial cells (d), endothelial cells (e), and leukocytes (f). (g-j) The heatmaps show the relative gene expression levels for the indicated marker genes for the indicated stromal (g), epithelial (h), endothelial (i), and leukocyte (j) cell types.

To increase statistical power and validate our results, we integrated our dataset with two large publicly available single cell RNA-seq datasets (Fig. 2a, b; Reyfman et al^17^ - Chicago cohort: ILD n=9, controls n=8; Habermann et al^15^ - Nashville cohort: ILD n=20, controls n=10). We generated a data manifold (as described in methods) that represents gene expression profiles of 233.638 single cells from 63 human individuals (ILD n=32, controls n=31). Cell type identities were then annotated manually as described above (Fig. 2c; Table S2). Next, we performed differential gene expression analysis between endstage lung fibrosis and control donors integrating all three study cohorts stratified by cell types as described in methods (see Table S3 for a full list of differential gene expression for health status stratified by cell type). Gene expression changes in disease were more similar within the epithelial, mesenchymal and leukocyte lineages (Fig. 2d), and showed very good agreement between the three independent patient cohorts (Fig. 2e-h).

**Figure 2.**
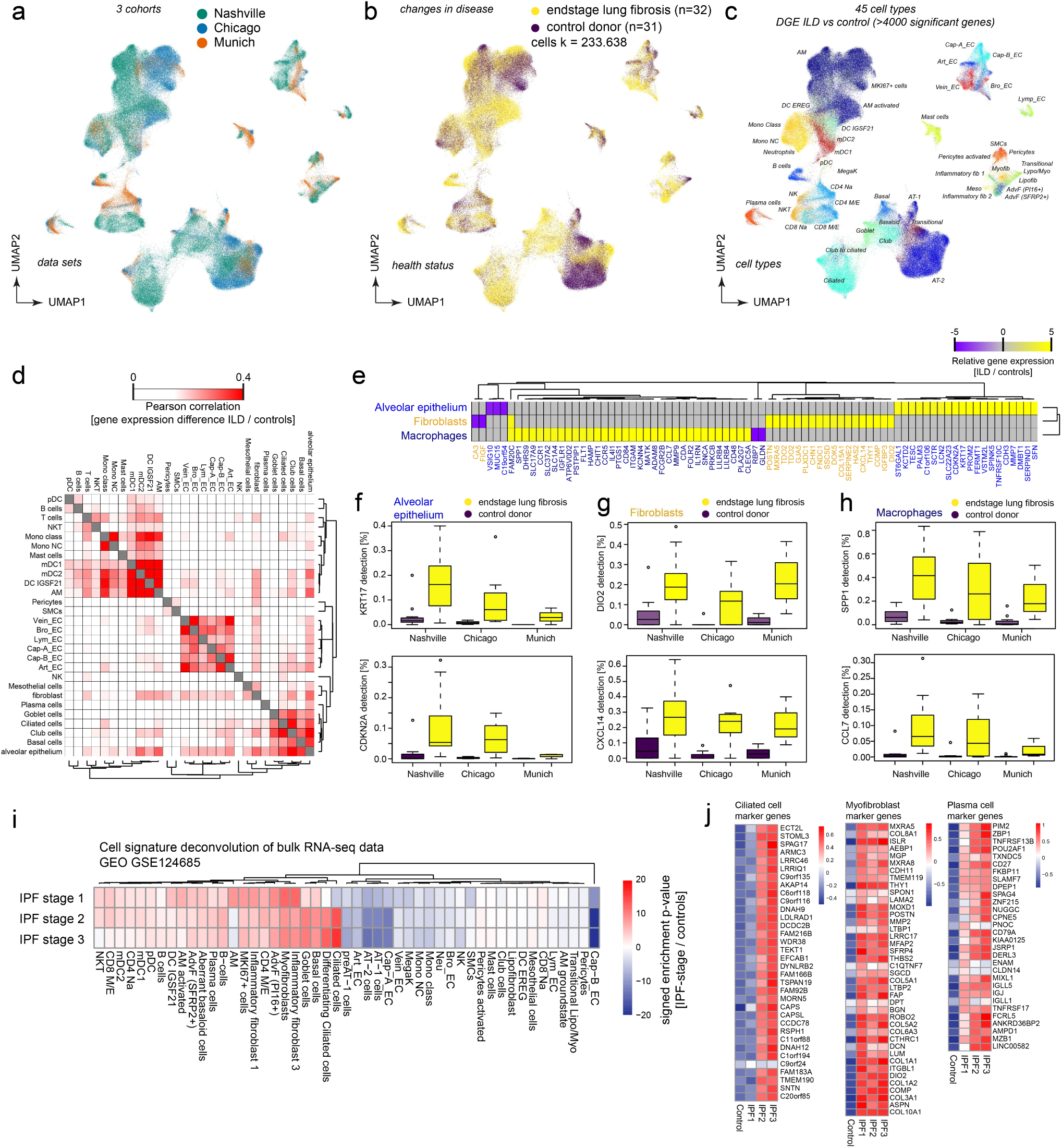
Multi-cohort single cell data reveals transcriptional changes in >40 cell types and altered cell type frequencies in disease progression. (a-c) Dimension reduced single cell transcriptomic data is visualized through Uniform Manifold Approximation and Projection (UMAP). The color code illustrates the patient cohort (a) disease status (b) and cell type identity (c). (d) Differential gene expression between endstage lung disease patients and controls across cohorts was compared for the indicated cell identities. The color code illustrates similarities of gene expression changes calculated by Pearson correlation of the t-value coefficient, which represents differences in likelihood of detection for any gene between health and disease. (e) The heatmap illustrates the top 79 genes differentially expressed in the indicated cell identities. (f-h) The box plots illustrate differences in mRNA detection for the indicated genes between tissues from control donors and fibrosis patients in f) alveolar epithelial cells, (g) fibroblasts, and (h) macrophages. (i) The heatmap shows changes of our cell type signatures in published bulk RNA-seq data (GEO GSE124685) across different histopathological stages that represent increasing extent of fibrosis from stage 1-3, as determined by quantitative micro-CT imaging and tissue histology^22^. (j) The heatmaps show z-scores for the individual marker genes of the indicated cell types across IPF stages and controls.

To leverage the power of bulk RNA-seq data archived in public databases, we used our ILD single cell atlas to determine possible cell type frequency changes in such datasets. A recent study used quantitative microCT imaging and tissue histology on biopsies to stratify lung tissue of idiopathic pulmonary fibrosis (IPF) patients into different stages marked by increasing extent of fibrotic remodeling (lower alveolar surface density and higher collagen content)^22^. Thus, the RNA-seq profiles of these staged patient samples presumably depict disease progression within patients. We calculated enrichment of our cell state signatures across the three stages of IPF progression and surprisingly found significant changes of many cell types already in early stage IPF 1 (Fig. 2i). This included the myofibroblast signature that was clearly upregulated early in progression. Other cell signatures, such as the plasma cells, showed a gradual increase from IPF1 to IPF3, while for instance the increase in ciliated cell frequency was observed only from IPF stage 2 onwards (Fig. 2j).

In summary, we generated an integrated and consistent lung cell atlas across three independent patient cohorts. Differential gene expression in lung fibrosis was robustly replicated and validated across cohorts and thus will serve as a powerful resource to investigators studying ILD pathogenesis and progression, e.g. for the dissecting of bulk RNA-seq profiles as demonstrated.

### Human lung bronchoalveolar lavage fluid proteomes reflect changes in disease activity

Transcriptional changes are not always correlated with protein abundance, in particular if proteins are secreted^12^. Some of the cell state changes described by our single cell analysis may be reflected in the proteomic composition of the luminal epithelial lining fluid (ELF) of the lung, which is accessible for sampling during bronchoscopic examination of patients. Here, we used state of the art mass spectrometry for in depth analysis of ELF proteins sampled from cell free bronchoalveloar lavage fluid (BALF) of a large ILD and non-ILD patient cohort (Fig. 3a). We have measured BALF proteomes from eight groups of patients that were diagnosed with different forms of ILD (Fig. 3b), including patients with idiopathic pulmonary fibrosis (IPF, n=16), hypersensitivity pneumonitis (HP/EAA, n=8), cryptogenic organizing pneumonia (COP, n=11), idiopathic non-specific interstitial pneumonia (NSIP, n=10), smoking associated respiratory bronchiolitis ILD (RB-ILD, n=3), Sarcoidosis (n=22), unclassifiable ILDs (other ILDs, n=25), and also included non-ILD controls (non-ILD, n=29) (see supplementary Table S4 for clinical features of the ILD cohort). Of note, the majority of lavage fluids from patients in this cohort was collected during evaluation of initial diagnosis of ILD, and thus rather represents early disease. From 124 patients (95 ILD and 29 non-ILD) that passed quality control criteria, we quantified a median number of 835 proteins per individual patient, resulting in a total of 1513 unique proteins that were quantified in at least 20 patients (Fig. 3b and Table S5). This is a very good depth of analysis given that BALF is difficult to analyze by mass spectrometry because of the high dynamic range of protein copy numbers present.

**Figure 3.**
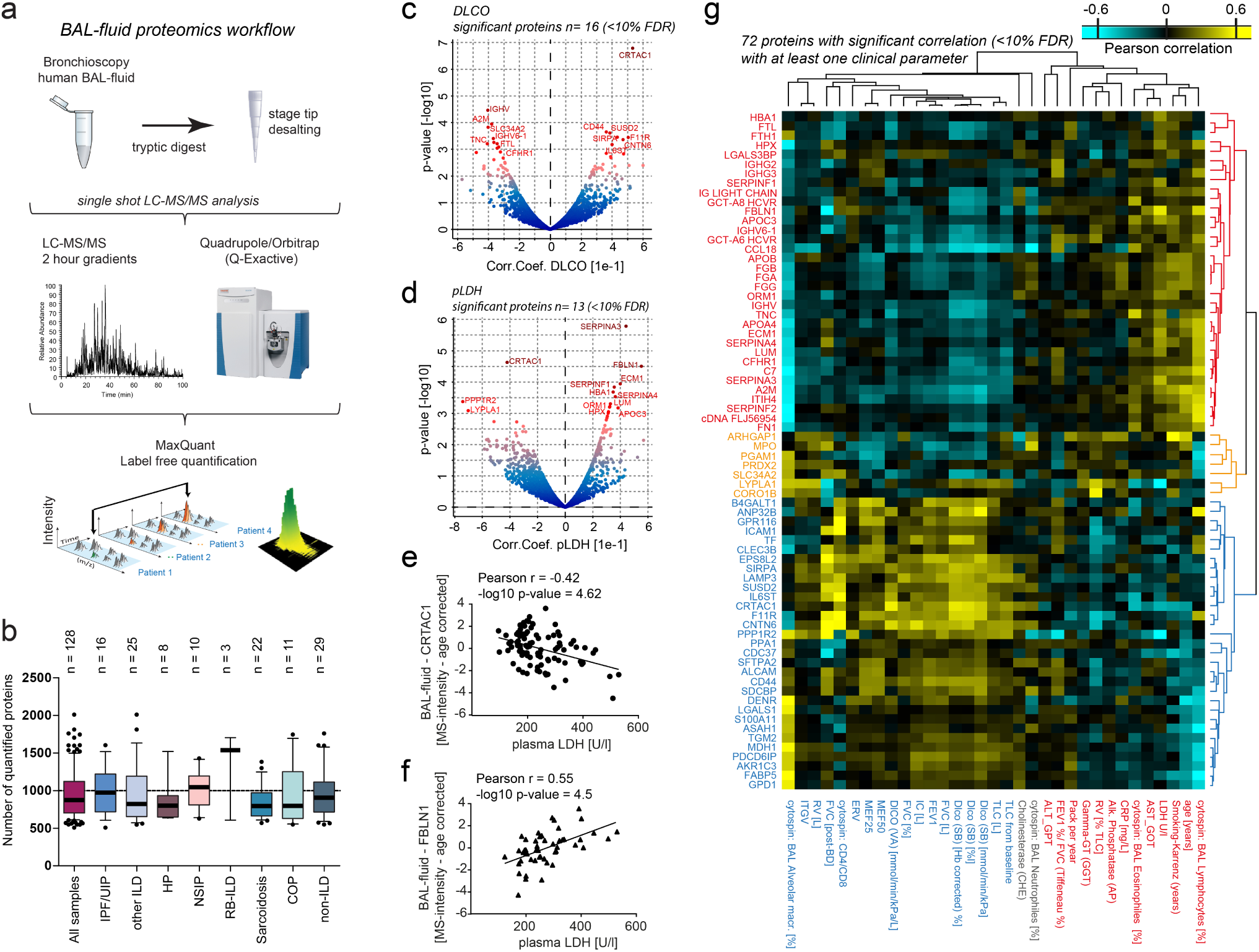
Human lung bronchoalveolar lavage fluid proteome changes correlate with clinical parameters. (a) Proteomics workflow. (b) The boxplots show the number of proteins quantified (y-axis) across various diagnoses (x-axis). The mean and 10-90 percentiles are shown. (c, d) The volcano plot shows Pearson correlation values (x-axis) and the –log10 p-value (y-axis) for BALF protein abundance and (c) DLCO and (d) plasma LDH (pLDH) values, respectively. (e, f) The scatter plots show (e) the negative correlation of CRTAC1 abundance in BALF (MS-intensity) with plasma LDH levels, and (f) the positive correlation of FBLN1 abundance in BALF (MS-intensity) with plasma LDH levels. (g) The Pearson correlation values for 72 significantly regulated proteins (<10% FDR) with indicated clinical parameters were grouped by hierarchical clustering into protein negatively and positively correlated with lung function (red and blue font respectively for negative and positive associations).

To better define the proteins that are true constituents of the ELF (rather than tissue leakage proteins), we made a quantitative comparison of BALF content and total tissue proteomes from 11 end-stage ILD tissue biopsies^6^. Proteins detected in both tissue and fluid proteomes were scored as either a ‘tissue leakage’ protein or true ‘epithelial lining fluid’ protein based on their enrichments in the respective compartments (Fig. S2a). Indeed, proteins specific for secretory epithelial cells such as Club and AT2 cells had a significantly higher ELF enrichment score as proteins specific to non-secretory AT1 cells. Similarly, we found that secreted proteins had a higher score than transmembrane proteins and cytoplasmic proteins (Fig. S2b). A Fisher’s exact test showed that 285 proteins with high coefficient of variation (CV) across patients (Fig. S2c) were significantly enriched for gene annotations such as ‘secreted’, ‘plasma lipoprotein’, ‘antimicrobial’, ‘nucleosome’, ‘intermediate filament’ and ‘extracellular matrix’ (Table S6), indicating that these categories are regulated across patient groups. Principal component analysis revealed clusters of patients with heterogeneous diagnosis that were either significantly enriched in complement, coagulation proteins and plasma lipid transport proteins, or showed higher levels of antimicrobial proteins and histones, pointing towards the involvement of an inflammatory response driven by neutrophil extracellular traps (NETs) in these patients (Fig. S2d, e). Correlation analysis of the protein expression profiles revealed that in fact many proteins were co-expressed across patients, revealing co-regulated protein modules that were enriched for distinct signatures, including macrophage specific proteins such as the Scavenger receptor cysteine-rich type 1 protein M130 (CD163) and the Complement C1q subcomponent subunit C (C1QC), wound healing factors, such as the ECM proteins Tenascin-C, Fibronectin, Collagen type 6 and Periostin, as well as lipid transport, complement and coagulation proteins such as Apolipoprotein B-100 and Complement component C7, or antimicrobial defense and neutrophil chemotaxis factors, including granulocyte specific proteins such as S100-A8, S100-A9, Cathelicidin antimicrobial peptide (CAMP) and Myeloperoxidase (MPO) (Fig. S2f).

To identify associations of these protein signatures with clinical parameters we performed correlation analysis with 33 individual clinical measurements per patient, including various lung function parameters and plasma LDH (Table S7 and Fig. S3). We identified biomarker signatures by Pearson correlation of the clinical parameters with proteins that were quantified in BALF in at least 20 patients (Table S2), revealing highly significant correlations of distinct sets sets of proteins with several lung function parameters, including Diffusing Capacity For Carbon Monoxide (DLCO) (Fig. 3c), or plasma levels for lactate dehydrogenase (pLDH) (Fig. 3d). LDH in blood plasma is routinely measured in the clinic as a biomarker for ongoing tissue damage. Most proteins that we found increased in BALF of patients with high pLDH were also associated with lower lung function (Fig. S4a), and top outliers remained significant after accounting for patient age (Fig. S4b-e). For instance, the cartilage acidic protein 1 (CRTAC1) showed very robust negative correlation (Fig. 3e), while the ECM protein Fibulin-1 (FBLN1) was positively correlated to pLDH (Fig. 3f). In total, we identified 72 proteins with significant correlation (<10% FDR) with at least one clinical parameter. Hierarchical clustering identified two main clusters of proteins based on correlations with lung function, demographics, laboratory values and cytospin results (Fig. 3g).

Because LDH is released during tissue damage and transpires to the blood, its levels in blood plasma are clinically used as a marker of common injuries and diseases such as heart failure. We hypothesized that BALF proteins with correlation to pLDH in human patients represent a lung injury signature. A comparison of the human pLDH signature with BALF proteomes from mice after bleomycin injury revealed similar outlier proteins across species including the injury marker Tenascin-C (Fig. S4f). Using 1D annotation enrichment analysis (see methods), we confirmed that the pLDH correlation revealed protein changes in human patient BALF proteomes that were highly similar to the ones that can be observed upon a defined acute lung injury in the bleomycin mouse model (Fig. S4g).

In summary, we analyzed BALF proteomes from 124 patients and correlated protein expression with an extensive set of clinical parameters, which represents, to the best of our knowledge, the most comprehensive characterization of human pulmonary epithelial lining fluid composition so far. We identified co-regulated protein modules that were associated with patients lung function and current injury status, suggesting that some of these protein signatures could be used to monitor acute or subclinical exacerbations of ILD patients.

### Correspondence of fluid proteins with transcriptional changes in specific cell types

Next, we aimed to explain quantitative changes in BALF protein signatures with the cell state changes analyzed by single cell RNA-seq. We first deconvoluted the diagnosis specific protein biomarker signatures in the BALF proteomes and evaluated the relative contribution of cell types/states. Mean intensity z-scores of proteins across different diagnostic groups were tested for enrichment of cell type specific transcriptional signatures (Fig. 4a).

**Figure 4.**
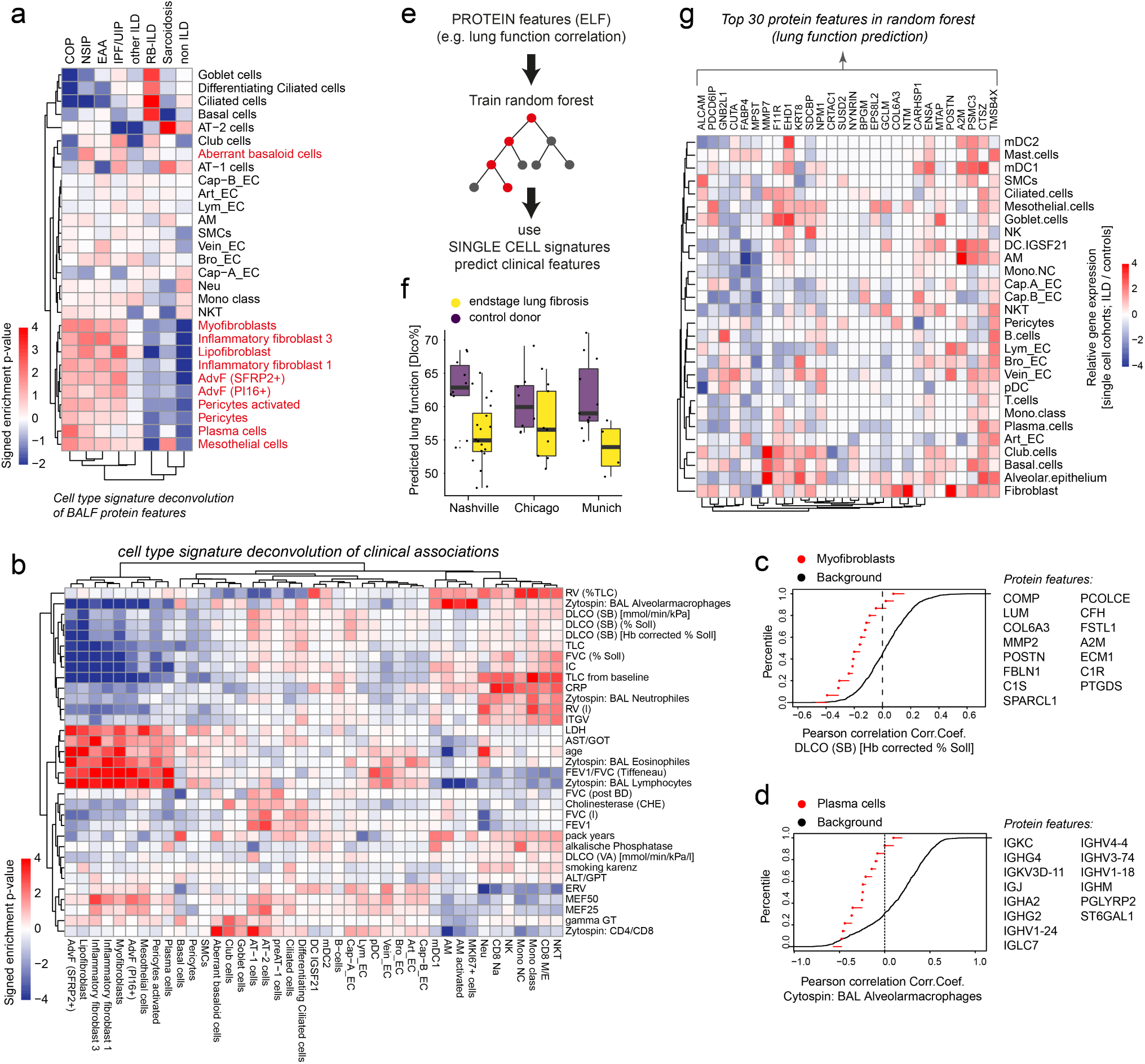
Protein signatures in BALF predict lung function decline and the corresponding cellular changes. (a) The heatmap shows relative contribution of cell types to the diagnosis specific protein biomarker signatures in epithelial lining fluid (ELF). (b) The heatmap shows relative contribution of cell types to the association of protein biomarker signatures in ELF with the indicated clinical parameters. (c, d) Empirical cumulative density plots show the distribution of correlation coefficients for (c) Myofibroblast markers (red points) with DLCO and (d) Plasma cell markers (red points) with % alveolar macrophages in BAL and the background proteins (black line). (e) The Pearson correlation of protein features in ELF was used to train a random forest algorithm. Training data was used on transcriptional signatures in single cell RNA-seq data to correctly predict reduced lung function in endstage lung fibrosis compared to control donors. (f) Box plots show predicted lung function changes (DLCO%) in the three single cell cell RNA-seq cohorts. (g) Top protein features in the random forest training data are shown with their relative gene expression changes in the different indicated cell types, illustrating cell type specific changes in lung fibrosis for these BALF biomarkers.

Markers of several pro-fibrogenic cell types including fibroblast subsets, pericytes, plasma cells and mesothelial cells were strongly increased in protein measurements of COP, NSIP, HP/EAA, and IPF compared to non-ILD controls, confirming the power of BALF proteomics to correctly score fibrogenic remodeling in the patients. Interestingly, RB-ILD and Sarcoidosis samples were similar to non-ILD controls for this signature, which is consistent with their distinct histopathology that does not involve strong interstitial fibrosis. While RB-ILD protein analysis featured very strong enrichment for proteins specific to airway basal, ciliated and goblet cells, the same airway protein signature was depleted in patients with COP, NSIP and HP/EAA but not IPF (Fig. 4a).

Similarly, we found strong associations of cell type/state signatures with the Pearson correlation of most clinical parameters with the protein measurements (Fig. 4b). For instance, the myofibroblast specific proteins quantified in patient BALF tended to be negatively correlated with lung function (DLCO) (Fig. 4c), and the number of alveolar macrophages in BAL cytospins tended to be negatively correlated with proteins (mostly antibodies) secreted by plasma cells into the ELF (Fig. 4c). Thus, deconvolution of protein measurements with cell type/state signatures revealed diagnosis and disease state specific biomarker fingerprints.

To test if we could successfully transfer information from the proteomics modality into the scRNAseq data modality we applied machine learning. A random forest was trained on the protein quantification data to predict lung function (DLCO) using a set of protein features which 1) showed high correlation with lung function (DLCO) and 2) and had the corresponding transcript detected in the scRNAseq data. Next, the trained model was applied to in silico bulk scRNAseq data with mRNA expression mapped to proteins (Fig. 4d), which then correctly predicted the direction of lung function changes in the three single cell RNA-seq cohorts (Fig. 4e). Indeed, the most important protein features of the random forest model appeared to be regulated in a cell type specific manner at the mRNA level (Fig. 4g).

In summary, our cross-modality analysis serves as proof of concept that cell state and frequency changes in diseased organs can transpire into predictive body fluid protein signatures that can be analyzed by mass spectrometry with high precision.

### Smoking induces highly persistent cell state changes in the lung

It is estimated that one hundred million deaths were caused by tobacco in the 20^th^ century^25^. Smoking is a major risk factor for six of the eight leading causes of deaths in the world, including respiratory and cardiovascular diseases, stroke and several malignant diseases^26^. Likewise, smoking might also play a major role in the genesis of some ILD entities. Some forms of ILD are directly smoking-related, such as RB-ILD and desquamative interstitial pneumonia (DIP)^27^, and smoking is also discussed to increase the risk for developing IPF^3^. We identified a robust smoking signature in the BALF proteomes of our ILD cohort. Clinical characteristics of patients included in the smoking analysis are shown in Table S8. A t-test between active smokers (n = 19) and never smokers (n = 36) identified 422 significantly regulated proteins (FDR<10%) (Fig. 5a), including AKR1B10 and the ubiquitin carboxyl-terminal hydrolase isozyme L1 (UCHL1), both of which have been shown to be elevated in epithelium of ‘healthy’ smokers^28,29^. The comparison of active smokers with ex-smokers (n = 49) identified 137 significantly regulated proteins (FDR<10%) (Fig. 5b). Of note, we also detected 36 significantly altered proteins (FDR<10%) in ex-smokers vs never smokers (Fig. 5c), indicating that some of the alterations induced by smoking are persisting after smoking cessation. Indeed, the fold changes between active/never and ex/never smokers were significantly correlated (Pearson r = 0.4785, -log10 p- value = 14.6536) (Fig. 5d). While surprisingly many changes in smokers persisted after cessation even over decades, we found a number of oxidoreductases, including AKR1B10, as highly specific markers of active smoking (Fig. 5d-f). The previously reported downregulation of the chemokine CCL18 in alveolar macrophages^30^, persisted after smoking cessation, suggesting that alveolar macrophages permanently change their phenotype (Fig. 5g, h). Also the scavenger receptor CD163 was downregulated in both active and ex-smokers compared to never smokers (Fig. 53i). The decline in CCL18 and CD163 was permanent and independent of time after cessation (Fig. 5h and 5j, respectively). In contrast to never smokers, the intensity distribution of CD163 was bimodal in smokers (Fig. 5k), indicating that only a subset of patients is affected in this manner by smoke exposure.

**Figure 5.**
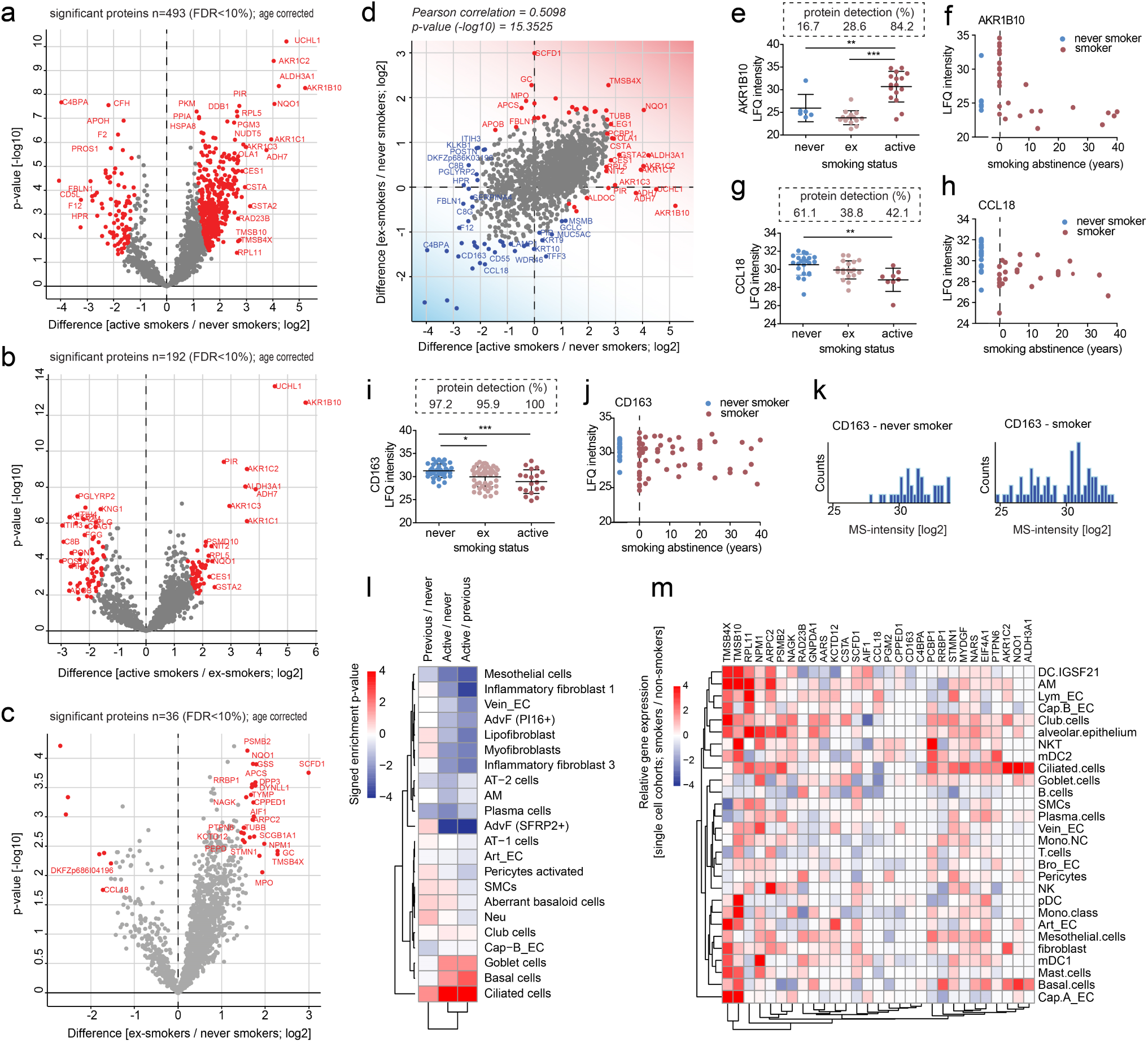
Smoking induces transient and persistent cellular changes reflected in BALF proteomes. (a-c) The volcano plots show significantly regulated proteins in BALF between (a) active smokers (n=19) and never-smokers (n=36), (b) active smokers and ex-smokers (n=49), and (c) ex-smokers and never-smokers. (d) The scatter plot compares the fold changes between active smokers vs. never-smokers (x-axis) and ex-smokers vs. never-smokers (y-axis). (e) AKR1B10 is significantly higher and more often detected in BAL fluid of active smokers in comparison to ex- (p<0.0001) and never smokers (p<0.0001). (f) Analysis of AKR1B10 and years of smoking abstinence show a reversible expression pattern of AKR1B10 after smoking cessation. (g) CCL18 is detected significantly more often in never vs ex-smokers (p=0.0417) and with significantly higher intensity in never vs active smokers. (h) The reduced levels of CCL18 do not go back to baseline after smoking cessation. (i) While CD163 was detected in almost all patients irrespectively of the smoking status, ex- and active smokers showed significantly lower protein levels in comparison to never smokers. (j) CD163 levels are changed in smokers and do not recover after smoking cessation. (k) The histograms show the distribution of MS-intensity values for CD163 in ELF across patients in never smokers (left panel) and smokers (ex- and active smokers – right panel). (l) The heatmap shows relative contribution of cell types to the specific protein biomarker signatures in epithelial lining fluid (ELF) of active, former and never smokers. (m) Top protein features in the smoking correlation data are shown with their relative gene expression changes in the different indicated cell types.

Next, we investigated the relative contribution of cell types/states to the association of protein biomarker signatures in ELF with patient smoking status (Fig. 5l). Proteins derived from airway epithelial cells were strongly enriched in ELF of smokers compared to non-smokers. Interestingly, the increase of basal and goblet cell derived proteins was reversible, while increased abundance of proteins derived from ciliated cells was persistent. Of note, most of the top proteins regulated with smoking status in ELF were also regulated on transcriptional level in the three single cell RNA-seq cohorts in at least one or many cell types (Fig. 5m; see Table S9 for a full list of differential gene expression for smoking status stratified by cell type).

In summary, we found that proteome changes in active smokers were partially persistent for decades after smoking cessation, indicating long lasting epigenetic alterations in airway epithelial cells and alveolar macrophages.

### Biomarkers of lung health in plasma proteomes

More than half of all clinical decisions are currently based on blood biomarker analysis^31^. To extend our analysis to the plasma proteome we made use of a recently established high throughput plasma proteomics workflow^31–33^ (Fig. 6a), and generated plasma proteomes from two independent cohorts of ILD patients (Munich, n=30 and Hannover, n=81; healthy age matched controls, n=30; see Table S10 for clinical characteristics and Table S11 for plasma protein quantifications). The Hannover cohort included more patients with better lung function on average, with samples taken mainly at time of initial diagnosis, while the Munich cohort contained patients closer to endstage disease. Thus, by construction, we did not expect a perfect match of these two cohorts. Upon correlation with forced vital capacity (FVC %) we identified a shared panel of proteins in both cohorts that were either positively or negatively associated with the lung function outcome (Fig. 6b).

**Figure 6.**
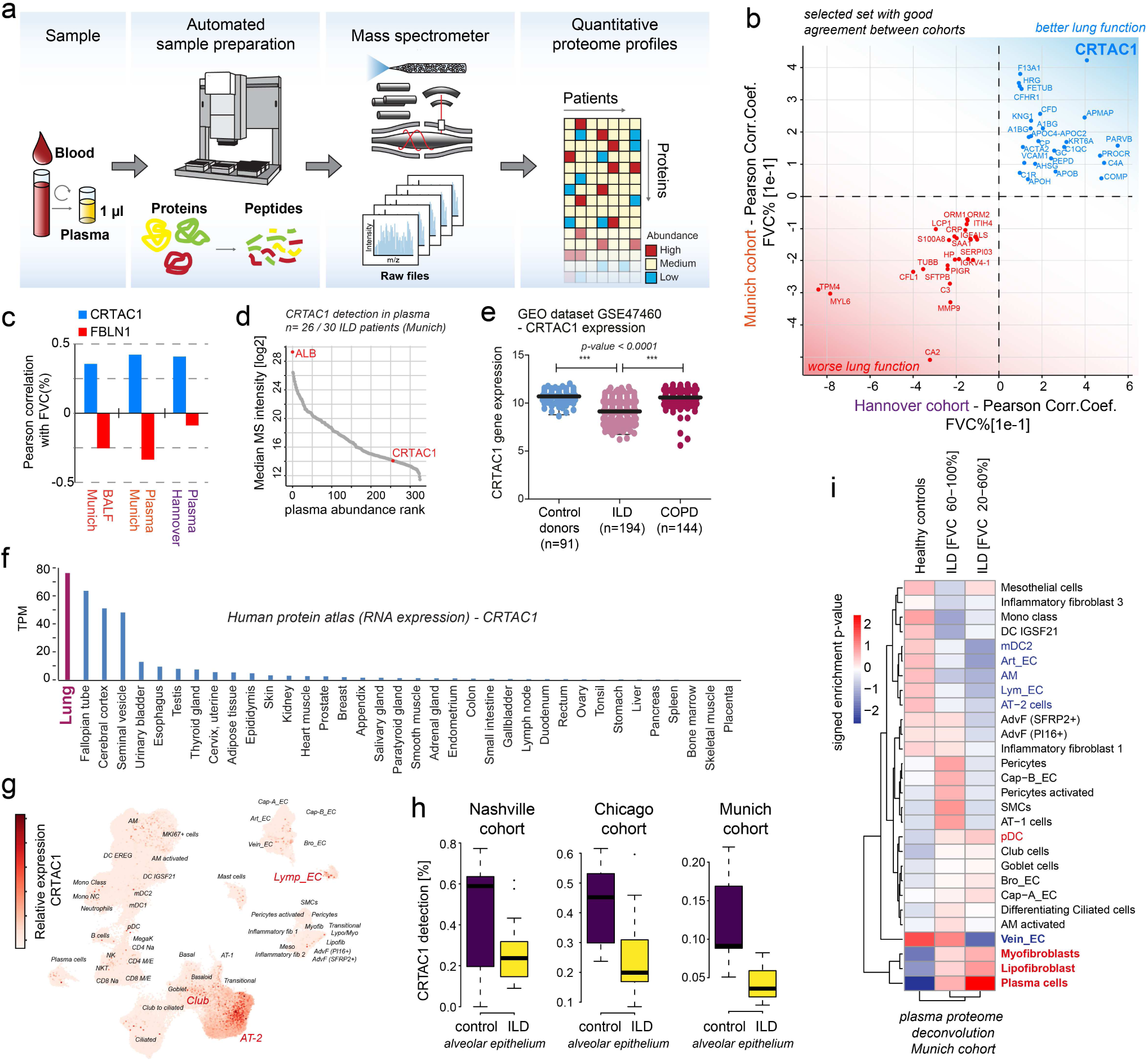
Multi-cohort plasma proteome analysis identifies the novel peripheral biomarker of lung health CRTAC1. (a) A high throughput experimental workflow for plasma proteomics^32^. (b) The indicated proteins were selected based on their common direction of correlation with patient lung function in two independent patient cohorts with distinct clinical characteristics. (c) The bar graph shows Pearson correlation coefficients of the indicated proteins with the lung function parameter forced vital capacity (FVC %) in the bronchoalveolar lavage fluid proteome cohort (BALF Munich), and two plasma proteome cohorts (plasma Munich, plasma Hannover). (d) All proteins quantified in plasma ranked by their abundance measured by mass spectrometry (MS-intensity). (e) Relative gene expression levels of *CRTAC1* in GSE47460. Dots represent average expression in tissue of individual patients. The line represents the mean. *CRTAC1* is significantly downregulated in ILD but not COPD patients (One-way ANOVA). (f) Relative expression level of *CRTAC1* across human organs. (g) UMAP embedded visualization of single cells colored by gene expression for *CRTAC1*, which is specifically expressed in alveolar type-2 (AT2), Club and lymphatic endothelial (Lymp_EC) cells. (h) The box plots illustrate differences in mRNA detection for *CRTAC1* in alveolar epithelial cells from control donors and fibrosis patients in the three indicated patient cohorts. (i) The heatmap shows the predicted relative contribution of lung cell types to the association of protein biomarker signatures in plasma with lung function (forced vital capacity - FVC). Patients were split in two groups, one with a mild decline in lung function [FVC 60-100%] and one with severe loss of lung function [FVC 20-60%] and compared to healthy age matched controls.

Most prominently we found the cartilage acidic protein 1 (CRTAC1) has higher levels in the plasma of patients with better lung function in both cohorts. CRTAC1 was also positively correlated with lung function in the BALF proteome analysis (Fig. 6c, Fig. 3), and was robustly detected by mass spectrometry in >80% of the plasma samples (Fig. 6d). Re-analysis of published bulk transcriptomes confirmed a highly significant downregulation of *CRTAC1 mRNA* in the lung of ILD patients compared to healthy controls and COPD patients (Fig. 6e). On whole body level the mRNA expression of *CRTAC1* is highest in the lung (Fig. 6g), and our single cell atlas reveals specific expression in lung lymphathic endothelium, airway club cells, and most prominently in alveolar type-2 epithelial cells (Fig. 6g). Expression of *CRTAC1* in alveolar epithelial cells was consistently downregulated in ILD samples compared to controls in all three patient cohorts analyzed by single cell RNA-seq (Fig. 6h).

Finally, we investigated the relative contribution of cell types/states to the protein biomarker signatures in plasma that correlated with lung function (Fig. 6i). We divided the patients into two groups representing mild and severe disease based on lung function (FVC %) and compared these two groups with healthy controls. We observed a gradual increase of proteins potentially derived from lung fibroblast subsets, plasma cells and pDCs and a gradual reduction of proteins potentially derived from lung endothelial cells, alveolar macrophages, AT2 cells, and mDC2 (Fig. 6i).

We conclude that plasma proteomes harbor protein biomarker signatures that report lung health, with CRTAC1 as a particular robust example that can be monitored with mass spectrometry workflows.

## Discussion

The field of single cell genomics has rapidly evolved and with the increasing availability of cell atlases is now moving towards the mechanistic characterization of pathogenesis and disease progression. We can conceptualize inter-individual variance within patient cohorts with a model in which patients at different stages of a disease progression trajectory will have their cells and tissues in different characteristic states. Body fluids potentially contain a composite representation of these disease stage specific differences in the form of proteins and possibly cell free DNA. We must deconvolute these composite signatures in order to make predictions about cell and tissue level changes in the patient. In this work we show a first proof of concept for this idea. We envision that training machine learning algorithms with large datasets of matched single cell genomic and fluid proteomic or sequencing readouts will enable new automated tools for clinical decision making^34^ and drug monitoring^35^.

In idiopathic pulmonary fibrosis (IPF), the most common form of lung fibrosis, the progressive replacement of lung parenchyma with scar tissue leads to respiratory failure with a median survival time of 2-4 years after diagnosis. Current models of disease pathogenesis propose that a combination of repetitive (micro)injuries to susceptible alveolar epithelial cells (AEC) with an aberrant repair response causes pathological interactions of AEC with fibroblasts and subsequent accumulation of scar tissue^36^. Human genetic data and preclinical models show that epithelial injury possibly caused by environmental exposures, can drive subsequent fibrosis, with a combination of genetic predisposition and aging thought to be related to the failed regenerative response in IPF. Using correlation of ELF protein abundance with levels of LDH in patient plasma we identified a human lung injury signature that we validated in a mouse model of acute lung injury. The protein injury signature contained known biomarkers of IPF, including MMP7 and CCL18, correlated with lung function decline and featured an increase of proteins derived from various cell types including fibroblast subsets, pericytes, plasma cells and mesothelial cells, as well as reduced levels of proteins derived from several other cell types including alveolar macrophages, airway and alveolar epithelial and endothelial cells. BALF and plasma levels of CRTAC1 were robustly correlated with lung function and pLDH inferred injury status and we show that the highest expression of CRTAC1 in the human body is found in AT2 cells. The function of CRTAC1 in the lung is unknown. Interestingly, the levels of CRTAC1 in isolated human AT2 cells increase upon differentiation with glucocorticoids^37^, which are known to be essential for alveolar maturation in lung development^38^. We propose that the downregulation of CRTAC1 in AT2 of ILD patients may hint at currently uncharacterized changes in glucocorticoid signaling in these cells.

ILD patients experience highly diverse clinical courses, with progression often accelerated due to acute exacerbations (AE), associated with a high mortality^39^. Efforts have been made to find predictive biomarkers for AE and disease outcome^20,40^. For instance elevated serum levels of AT2 derived SP-A and SP-D are associated with an increased risk of mortality in IPF^40–42^. Nevertheless, such biomarkers are currently not clinically established and often it is unclear which cellular changes they represent. While early detection of AE is currently of major interest, there are also patients who present with clinical worsening without meeting the criteria for AE. A daily home spirometry study resulted in highly diverse lung function trajectories in IPF^43^, suggesting that lung function diversity could also reflect different stages after epithelial lung injury with phases of decreased lung function (potentially being a phase of subclinical injury/exacerbation) followed by phases of slightly increased lung function (potentially being a phase of successful tissue repair). Therefore, we propose that the human lung injury signature discovered in this study can instruct the design of follow up studies that evaluate its use in monitoring acute or subclinical exacerbations of ILD patients.

We identified a surprisingly large set of changes between ex-smokers and never smokers, some of which persisted even after decades of smoking cessation, indicating permanent epigenetic alterations. Indeed, signatures of smoking induced changes in genome-wide DNA methylation signatures have been shown to persist over many years^44^. Deconvolution of BALF proteomes with cell type signatures showed a strong increase in contributions from airway club and goblet cells as well as ciliated cells in active smokers, which was most prominent in smoking associated RB-ILD patients. Interestingly, the increase in club/goblet cell derived proteins, which likely stems from smoking induced goblet cell hyperplasia^45^, was fully reversible upon smoking cessation, while the number of proteins derived from ciliated cells remained upregulated in ex-smokers. This indicates a permanent shift in airway epithelial cell composition in ex-smokers. Top hits in the differential gene expression analysis between smokers and non-smokers in the three single cell cohorts also showed a bias towards airway epithelial cells, however not surprisingly also alveolar macrophages (AM) were severely affected. For instance, we observed a negative correlation of AM derived CCL18 with lung function and also an irreversible downregulation of the protein in BALF once the patient ever smoked. CCL18 has been reported to be elevated in the serum and BALF in patients with interstitial lung disease^46^. A pooled post-hoc analysis of the CAPACITY and ASCEND studies identified CCL-18 as the most robust blood marker for disease progression in IPF^20^. Given our observation that CCL18 levels are permanently reduced in ex-smokers it will thus be interesting to re-evaluate previous reports on CCL18 association with IPF progression with respect to smoking history.

Our machine learning analysis demonstrates that correspondence of fluid proteomes and single cell transcriptomes can be used to correctly predict the direction of lung function changes across modalities, indicating that further development of this concept will contribute to future clinical decision making. Our work has several limitations that prohibit to fully complete this task. Since the cross-modal analysis was done on non-matched patient cohorts it is currently difficult to assess the specificity of fluid proteome signatures for tissue level cell state changes, and to go beyond associative signatures. Thus, carefully designed longitudinal multi-modal analysis of animal models and patient cohorts will be required to train machine learning algorithms, in particular causal inference models, for future applications in predictive personalized medicine.

## Supplementary figures

**Figure S1.**
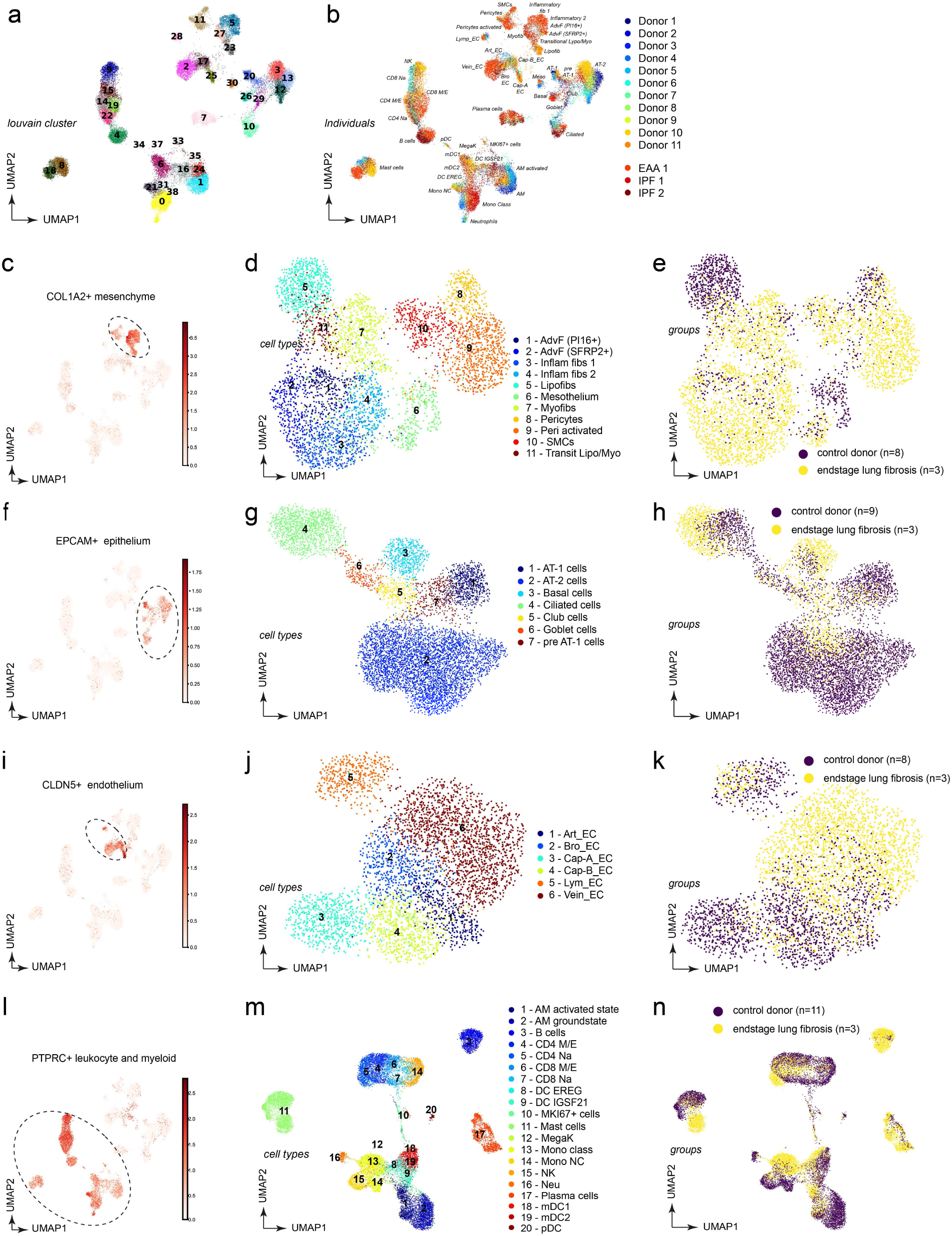
Clustering analysis and cell type annotation reveals 45 distinct cell type identities in human lung parenchyma. (a) UMAP embedding colored by Louvain clusters demonstrates separation of cells into major lineages. (b) UMAP embedding displays identified cell types, colored by individual patients. (c, f, i, l) The whole lung parenchymal dataset was split into subsets for (c) COL1A2+ mesenchymal cells, EPCAM+ epithelial cells (f), CLDN5+ endothelial cells (i) and CD45+ (gene name PTPRC) leukocytes (l). (d, g, j, m) New UMAP embeddings of the subsets demonstrate separation of cluster identities that allows for identification of cell states. (e, h, k, n) Cells colored in disease groups show origin of identified cell states.

**Figure S2.**
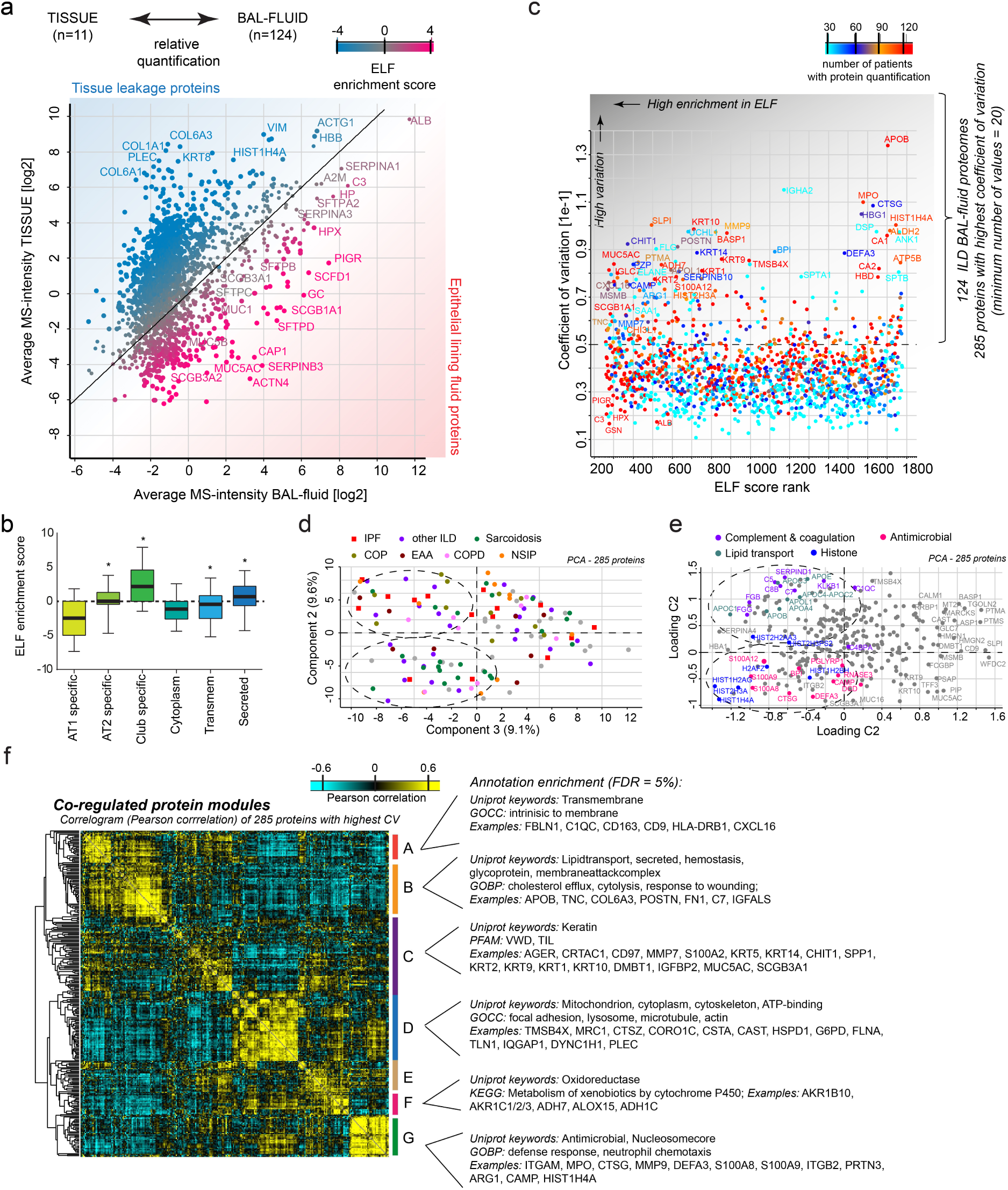
Heterogeneity of the epithelial lining fluid proteome in a large interstitial lung disease patient cohort. (a) Comparison of BAL fluid proteome (n=128) and ILD lung tissue proteome (n=11) allows the separation of true constituents of the epithelial lining fluid from tissue leakage proteins and identifies 199 proteins with significant enrichment in the BAL-fluid proteome. The color code shows the relative enrichment of proteins in fluid versus tissue (ELF score). (b) The box plot shows distributions of ELF enrichment scores for the indicated gene categories. (c) Identification of 285 proteins with highest coefficient of variation and heterogeneity between patients enriched in human BAL-fluid. (d, e) Principal component analysis reveals groups of patients with similar enrichment for the indicated gene categories, irrespective of the indicated clinical diagnosis. (f) The correlogram shows the Pearson correlation of the 285 proteins with highest CV across individual patients. Enriched gene categories and examples of co-regulated proteins are shown.

**Figure S3.**
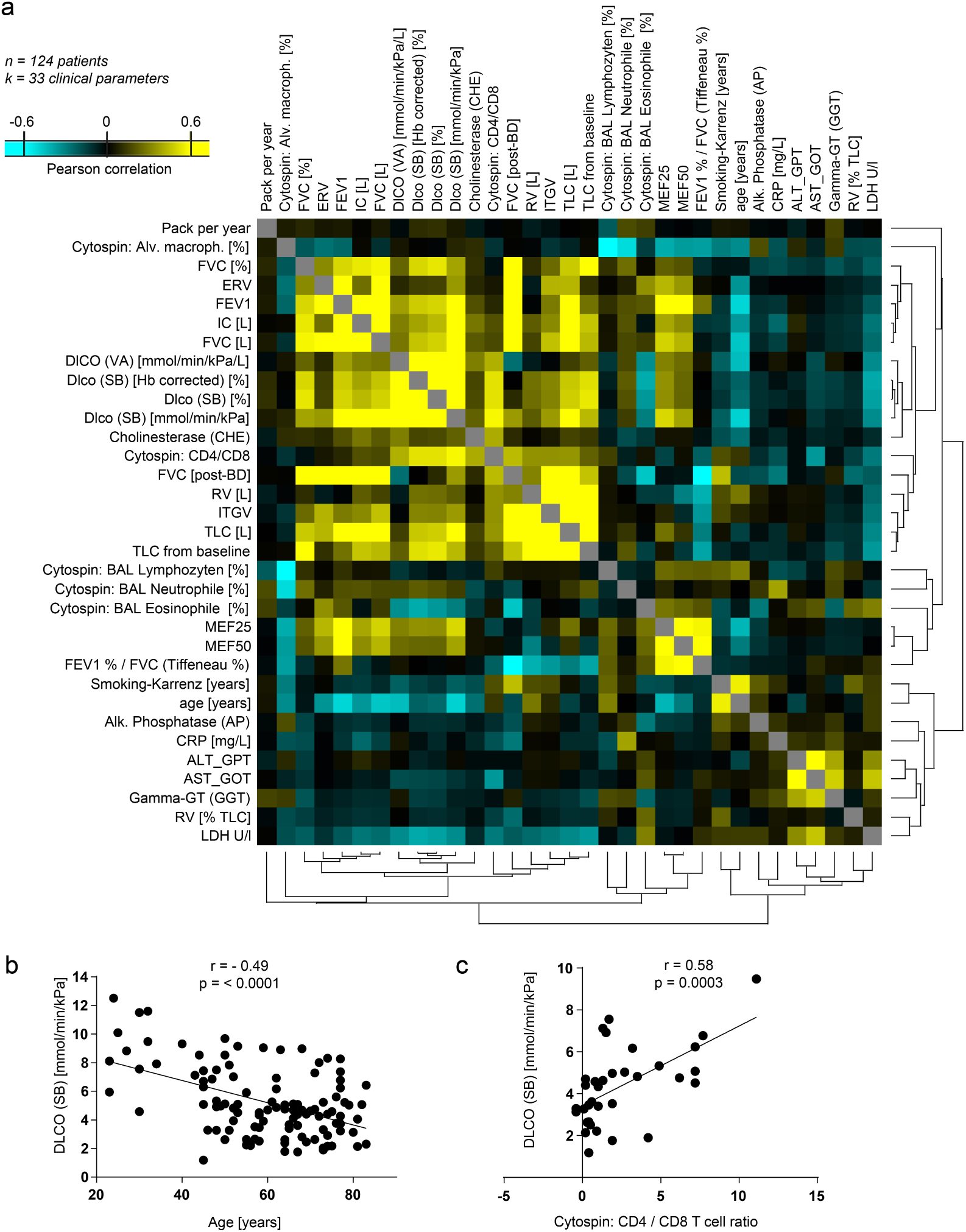
Correlation patterns between 33 clinical parameters in an ILD cohort. (a) Pairwise Pearson correlation values of 33 clinical parameters were grouped by hierarchical cluster analysis. (b) DLCO shows negative correlation with age in the study cohort (p<0.0001). (c) Positive correlation of CD4/CD8 ratio in BAL fluids with DLCO.

**Figure S4.**
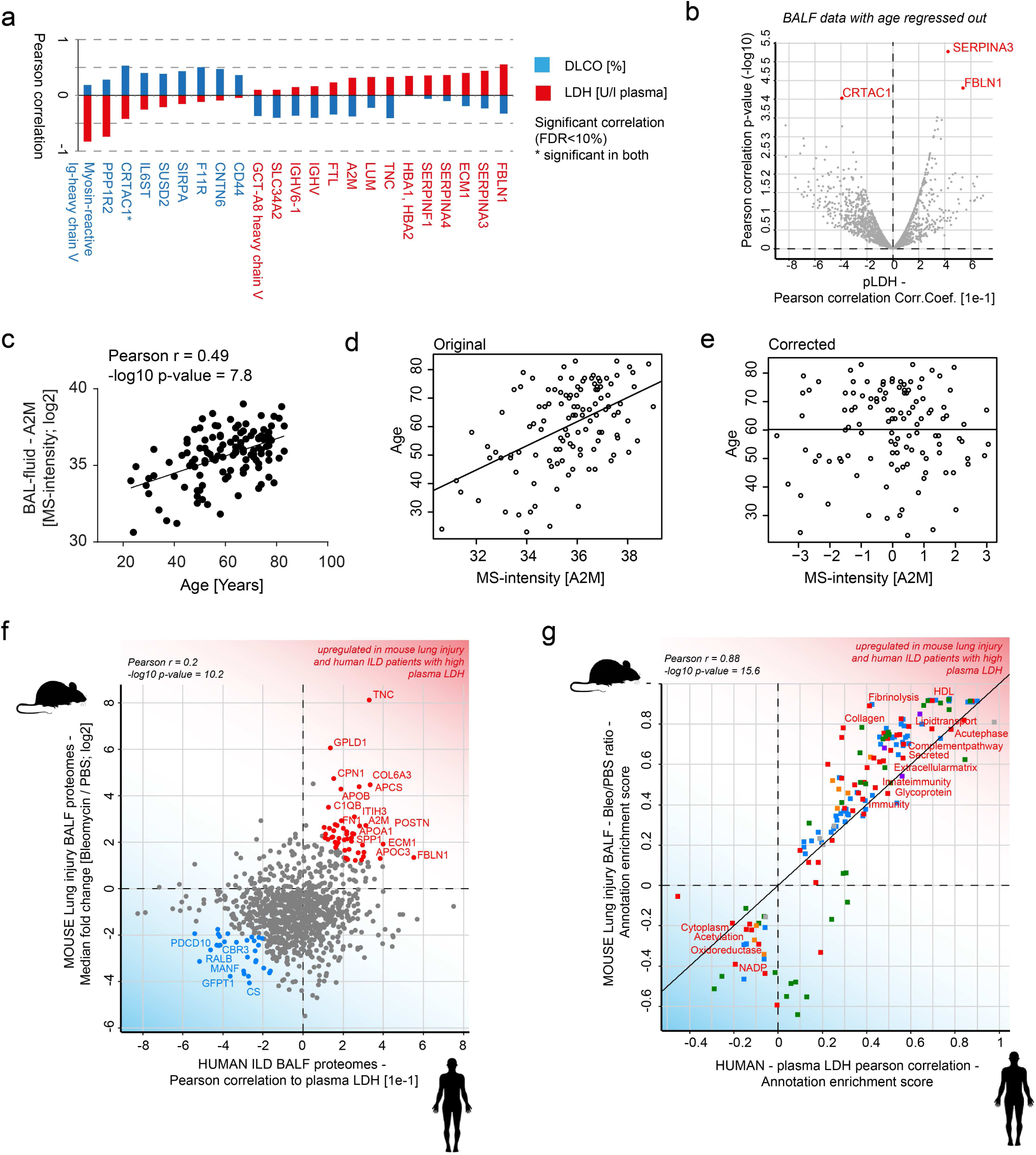
BALF proteins correlating with plasma LDH represent a human lung injury signature. (a) The bar graph shows the Pearson correlation values of the indicated proteins for DLCO [%] (blue) and plasma LDH (red). (c) The scatter plot shows significant correlation of alpha 2 macroglobulin (A2M) abundance (MS-intensity) in BALF with patient age. (d, e) Original (d) and age-corrected (e) correlation of A2M in BAL fluids with age. (f) The scatter plot shows the Pearson correlation of individual proteins from the human BAL fluid proteome with plasma LDH (x-axis) and the fold changes (y-axis) of the orthologous proteins in mouse lung after bleomycin injury^47^. (g) The annotation enrichment score shows a common upregulation of gene categories like acute phase, ECM, complement and innate immunity in the BAL fluids of bleomycin mice and human ILD with high plasma LDH.

## Methods

### Data availability

Count tables of the Munich single cell cohort as well as all custom analysis code can be accessed at https://github.com/theislab/2020_Mayr. Proteome raw data and MaxQuant processing tables can be downloaded from the PRIDE repository under the accession number PXD017145 (BALF) and PXD017210 (plasma).

### Human samples

Human samples of the Munich cohorts (tissue, BAL fluid and plasma) were obtained from the BioArchive of the Comprehensive Pneumology Center Munich (CPC-M). All patients gave written informed consent and the study was approved by the local ethics committee of the Ludwig-Maximilians University of Munich, Germany (EK 333-10 and 382-10). Plasma samples of the Hannover cohort were obtained as part of a cooperation of the German Centers for Lung Research (DZL). Patients gave written informed consent to the DZL broad- consent form and the study was approved by the local ethics committee of the Medizinische Hochschule Hannover (2923-2015). Lung tissues used as healthy controls for single cell analysis were tumor free, uninvolved lung tissue freshly obtained during tumor resections performed at the lung specialist clinic “Asklepios Fachkliniken Munich-Gauting”. ILD lung tissue for single cell analysis was freshly obtained after lung transplantation at the University Hospital Munich. BAL fluid samples of the BAL fluid cohort and matched plasma samples were collected at the lung specialist clinic and included mainly first ILD evaluations. Plasma samples from an independent ILD cohort were obtained from the university hospital of the Ludwigs Maximilian University Munich from patients seen in the ILD outpatient clinic during routine visits or in the inpatient unit during evaluation for lung transplantation.

The diagnosis of IPF was made in accordance with the current guidelines^48^. All ILD diagnosis were made according to international guidelines and established criteria. Non-ILD patients of the BAL fluid cohort included patients who underwent BAL due to evaluation of asthma, COPD, lung cancer, hemoptysis or chronic cough.

For transport from the surgeon to the laboratory, lung tissue samples for single cell analysis were stored in ice-cold DMEM-F12 media and packed in thermo stable boxes. Tissue was processed with a maximum delay of 2 h after surgery. On delivery to the lab, tissue samples were assessed visually for qualification for the study.

### Lung tissue processing

Lung tissue was processed as previously described^10^. Briefly, around 1.5 g of tissue per sample was manually homogenized into smaller pieces (∼0.5 mm^2^ per piece). Before tissue digestion, lung homogenates were cleared by washing excessive blood through a 40 μm strainer with of ice-cold PBS. The tissue was transferred into enzyme mix consisting of dispase, collagenase, elastase, and DNase for mild enzymatic digestion for 1 h at 37 °C while shaking. Enzyme activity was inhibited by adding PBS supplemented with 10% FCS. Dissociated cells in suspension were passed through a 70 μm strainer and pelleted. The cell pellet was resuspended in red blood cell lysis buffer and incubated shortly at room temperature to lyse remaining red blood cells. After incubation, PBS supplemented with 10% FCS was added to the suspension and the cells were pelleted. The cells were taken up in PBS supplemented with 10% FCS, counted using a Neubauer chamber, and critically assessed for single-cell separation and viability. Two-hundred and fifty thousand cells were aliquoted in PBS supplemented with 0.04% of bovine serum albumin and loaded for Drop-seq at a final concentration of 100 cells μl−1.

### Single cell sequencing using Dropseq

Drop-seq experiments were performed largely as described previously^49^ with few adaptations during the single-cell library preparation^12,50^. Briefly, using a microfluidic polydimethylsiloxane device (Nanoshift), single cells from the lung cell suspension were co-encapsulated in droplets with barcoded beads (ChemGenes). Droplet emulsions were collected for 15 min each before droplet breakage was performed using perfluorooctanol (Sigma-Aldrich). After breakage, beads were collected and the hybridized mRNA transcripts reverse transcribed (Maxima RT, Thermo Fisher). Unused primers were removed by the addition of exonuclease I (New England Biolabs). Beads were washed, counted, and aliquoted for pre-amplification with 12 PCR cycles (primers, chemistry, and cycle conditions identical to those previously described). PCR products were pooled and purified twice by 0.6x clean-up beads (CleanNA). Before tagmentation, cDNA samples were loaded on a DNA High Sensitivity Chip on the 2100 Bioanalyzer (Agilent) to ensure transcript integrity, purity, and amount. For each sample, 1 ng of pre-amplified cDNA from an estimated 1,000 cells was tagmented by Nextera XT (Illumina) with a custom P5 primer (Integrated DNA Technologies). Single-cell libraries were sequenced in a 100 bp paired-end run on the Illumina HiSeq4000 using 0.2 nM denatured sample and 5% PhiX spike-in. For priming of read 1, 0.5 μM Read1CustSeqB (primer sequence: GCCTGTCCGCGGAAGCAGTGGTATCAACGCAGAGTAC) was used.

### Processing of single cell data from Munich

For the single cell data of human patients form the Munich cohort, the Dropseq computational pipeline was used (version 2.0) as previously described^51^. Briefly, STAR (version 2.5.2a) was used for mapping^52^. Reads were aligned to the hg19 reference genome (GSE63269). For barcode filtering, we excluded barcodes with less than 200 detected genes. For further filtering we kept the top barcodes based on UMI count per cell, guided by the number of estimated cells per sample. As we observed a certain degree of ambient RNA bias, we applied SoupX^53^ to lessen this effect. The pCut parameter was set to 0.3 within each sample before merging the count matrices together. The merged expression table was then pre-processed further. A high proportion (> 10%) of transcript counts derived from mitochondria-encoded genes may indicate low cell quality, and we removed these unqualified cells from downstream analysis. Cells with a high number of UMI counts may represent doublets, thus only cells with less than 4000 UMIs were used in downstream analysis. Genes were only considered if they were expressed in at least 3 cells in the data set^52^.

### Analysis of single cell data from Munich

The downstream analysis of the Munich single cell data was performed using the Scanpy Package^54^, a python package for the exploration of single-cell RNA-seq data. Following the common procedure, the expression matrices were normalized using *scran*’s^55^ normalization based on size factors which are calculated and used to scale the counts in each cell. Next log transformation was used via scanpy’s pp.log1p(). Highly variable genes were selected as follows. First the function pp.highly_variable_genes() was executed for each sample separately, returning the top 4,000 variable genes per sample. Next, we only considered a gene as variable if it was labelled as such in at least 2 samples, resulting in a total of 15,096 genes which were further used for the principal component analysis. In an additional step to mitigate the effects of unwanted sources of cell- to-cell variation, we regressed out the number of UMI counts, percentage of mitochondrial DNA and the calculated cell cycle score using the function pp.regress_out().

For visualizing the whole Munich data set, the UMAP was generated using 50 components as input for scanpy’s tl.umap() with number of neighbors set to 10 and min_dist parameter to 0.4. To better align the data of the different patients and to account for possible batch effects we used the python package bbknn() (batch balanced k nearest neighbours)^56^ with the same number of components and neighbors. Louvain clustering was calculated with resolution 6. The whole lung parenchymal dataset was split into subsets for COL1A2+ mesenchymal cells, EPCAM+ epithelial cells, CLDN5+ endothelial cells and PTPRC+ leukocytes. New UMAP embeddings of these subsets were calculated until clear separation of cluster identities was achieved that allowed for identification of cell states by exploring the highest expressed markers per cluster explored via tl.rank_genes_groups() and manual assessment of known marker gene expression.

### Computational data integration of single cell data

To improve statistical power, to ensure generalization across cohorts and to achieve a more balanced ratio of diseased and healthy patients, our Munich single-cell RNA-seq data set was combined with the filtered count matrices from the Chicago cohort (Reyfman et al^17^) and the Nashville cohort (Habermann et al^15^).

Before combining these, the count matrices from Chicago and Nashville were processed separately. The normalization using scran and the log transformation of the two external datasets was performed as described for the Munich cohort. The effect of cell cycle, the percentage of mitochondrial reads and the number of UMI counts was regressed out cohort-wise as well.

For a first lighter batch correction we defined the list of variable genes in a way to decrease cohort specific effect as follows. For both the Nashville and the Chicago data we considered a gene as highly variable if it is labelled highly variable in at least 3 patients of the respective data set. Next, the preprocessed count matrices from the three data sets were merged and genes retained their highly variable status if they were highly variable in at least two of the three cohorts, resulting in 3,854 variable genes.

The concatenated object was scaled with scanpy’s pp.scale() function and the principal components were calculated using the defined variable genes. As a second batch correction we calculated the neighborhood graph using the bbknn package, defining the individual patients as batch key, 5 number of neighbors within batch and 40 components. As described for the Munich cohort, the whole combined object was subsetted and new embeddings were calculated in order to identify cell states.

### Differential gene expression analysis

To identify genes associated with ILD status in a cell-type specific manner we applied the following procedure. The R statistical software was used for the analysis (ref). Since the outcome of interest (ILD status) varies at the donor (n = 61) as opposed to the cell level (n = 233,638) we framed the analysis as a likelihood of detection problem across all donors. For each donor and cell-type combination we calculated the likelihood of detection for each gene as the average number of cells with more than one count. As the likelihood of detection represents a probability and is bounded between 0 and 1, values were square-root transformed. Next, we used multiple linear regression to model the probabilities of detection. The square-root transformed detection probability was used as the dependent variable and the ILD status, age or smoking status, as the explanatory variable accounting for the total number of UMI counts, total number of cells and study indicator as covariates. The resulting t and p-values for the coefficient describing the ILD status were used in downstream analysis.

### Cell type signature enrichment analysis

To infer cell type frequency changes from bulk transcriptomics or proteomics data we applied signature enrichment analysis. We defined cell type signatures as sets of genes with significant cell type specific expression as defined in Table S2. Next, we statistically evaluated enrichment of each signature in a ranked list of fold changes or correlation coefficients using the Kolmogorov-Smirnov test. The signed p-value score represents the -log10 p-value of the Kolmogorov-Smirnov test signed by the effect size. Negative and positive values represent depletion and enrichment of the given signature in the ranked list, respectively.

### Random forest prediction

To integrate scRNAseq with BALF data we used a random forest as implemented in the R *randomForest* package. First, BALF expression data was quantile normalized and scaled. Next, only features with an absolute correlation coefficient greater than 0.2 with lung function and present in the scRNAseq data were used to train a random forest to predict lung function. Then, *in silico* bulk scRNAseq was calculated by taking the mean expression count of each gene across all cells for all samples. Finally, the *in silico* bulk data was quantile normalized and scaled before feeding it into the trained model to predict lung function.

### Clinical parameters

For all patients included in the final analysis clinical information were collected at the time of BAL fluid procedure or when plasma was taken, respectively. Clinical parameters included demographics (age, gender, smoking status, pack years, smoking abstinence, lung function [forced vital capacity (FVC) (% pred.), FVC (l), FVC (post broncholysis), expiratory reserve volume (ERV), forced expiratory volume in 1 second (FEV1) (l), FEV1/FVC (%), inspiratory capacity (IC) (l), total lung capacity (TLC) (l), TLC from baseline, residual volume (RV), RV (%TLC), diffusing capacity of the lung for carbon monoxide (DLCO) (VA) (mmol/min/kPa/l), DLCO (SB) (% pred.), DLCO (SB) (Hb corrected, % pred.), DLCO (SB) (mmol/min/kPa), mean expiratory flow (MEF) 25, MEF50, intrathoracic gas volumen (ITGV)], laboratory values [cholinesterase, alkaline phosphatase, C-reactive protein, alanine-aminotransferase (ALT), aspartate-aminotransferase (AST), gamma-glutamyltransferase (GGT), LDH].

### BAL procedure

BALF was collected from 141 patients undergoing bronchoscopy from January 2013 until March 2016 at the Lungenfachklinik Gauting in Munich, Germany. Most of the patients underwent bronchoscopy due to ILD evaluation. BAL was performed with standard technique. In brief, 100 to 200 ml of sterile saline (0.9% NaCl) was instilled into the right middle lobe or the lingula in 20-ml injections which were each immediately aspirated. Cells of the BAL were analyzed by cytospin analysis. The remaining cell-free BAL fluid was immediately stored at -80°C and transferred to the BioArchive of the CPC-M. For mass spectrometry, only the cell-free BAL fluids were analyzed. Of the 141 patients only 124 passed quality control and were included in the analysis (95 ILD and 29 non-ILD).

### Mass spectrometry

The BAL fluid depleted from cells was subjected to mass spectrometry analysis. Proteins were precipitated from 300µl BAL fluid using 80% ice cold acetone, followed by reduction and alkylation of proteins and overnight digestion into peptides using Trypsin and LysC proteases (1:100) as previously described (Schiller et al, MSB 2015). Peptides were purified using stage-tips containing a Poly-styrene-divinylbenzene copolymer modified with sulfonic acid groups (SDB-RPS) material (3M, St. Paul, MN 55144-1000, USA) as previously described (Kulak et al, 2014). Approximately 2 μg of peptides were separated in four hour gradients on a 50-cm long (75-μm inner diameter) column packed in-house with ReproSil-Pur C18-AQ 1.9 μm resin (Dr. Maisch GmbH). Reverse-phase chromatography was performed with an EASY-nLC 1000 ultra-high pressure system (Thermo Fisher Scientific), which was coupled to a Q-Exactive Mass Spectrometer (Thermo Scientific). Peptides were loaded with buffer A (0.1% (v/v) formic acid) and eluted with a nonlinear 240-min gradient of 5–60% buffer B (0.1% (v/v) formic acid, 80% (v/v) acetonitrile) at a flow rate of 250 nl/min. After each gradient, the column was washed with 95% buffer B and re-equilibrated with buffer A. Column temperature was kept at 50 °C by an in-house designed oven with a Peltier element (Thakur et al, 2011) and operational parameters were monitored in real time by the SprayQC software (Scheltema & Mann, 2012). MS data were acquired with a shotgun proteomics method, where in each cycle a full scan, providing an overview of the full complement of isotope patterns visible at that particular time point, is follow by up-to ten data-dependent MS/MS scans on the most abundant not yet sequenced isotopes (top10 method) (Michalski et al, 2011a). Target value for the full scan MS spectra was 3 × 10^6^ charges in the 300−1,650 *m/z* range with a maximum injection time of 20 ms and a resolution of 70,000 at *m/z* 400. The resulting mass spectra were processed using the MaxQuant software (Cox and Mann, 2008), which enabled label free protein quantification (Tyanova et al., 2016).

Plasma samples were prepared with the Plasma Proteome Profiling Pipeline^57^ automated on an Agilent Bravo liquid handling platform. Briefly, plasma samples were diluted 1:10 in ddH2O and 10 µl were mixed with 10 µl PreOmics lysis buffer (P.O. 00001, PreOmics GmbH) for reduction of disulfide bridges, cysteine alkylation and protein denaturation at 95°C for 10 min. Trypsin and LysC were added at a ratio of 1:100 micrograms of enzyme to micrograms of protein after a 5 min cooling step at room temperature. Digestion was performed at 37 °C for 1 h. An amount of 20 µg of peptides was loaded on two 14-gauge StageTip plugs, followed by consecutive purification steps according to the PreOmics iST protocol (www.preomics.com). The StageTips were centrifuged using an in-house 3D-printed StageTip centrifugal device at 1500 g. The collected material was completely dried using a SpeedVac centrifuge at 60 °C (Eppendorf, Concentrator plus). Peptides were suspended in buffer A* (2% acetonitrile (v/v), 0.1% formic acid (v/v)) and shaking for 10 min at room temperature. Plasma peptides were measured using LC-MS instrumentation consisting of an Evosep One (Evosep), which was online coupled to a Q Exactive HF Orbitrap (Thermo Fisher Scientific). Peptides were separated on 15 cm capillary columns (ID: 150 µm; in-house packed into the pulled tip with ReproSil-Pur C18-AQ 1.9 µm resin (Dr. Maisch GmbH)). For each LC-MS/MS analysis about 0.5 µg peptides were loaded and separated using the Evosep 60 samples method. Column temperature was kept at 60 °C by an in-house-developed oven containing a Peltier element, and parameters were monitored in real time by the SprayQC software. MS data was acquired with data independent acquisition using a full scan at a resolution of 120,000 at m/z 200, followed by 22 MS/MS scans at a resolution of 30,000. MS raw files were analyzed by Spectronaut software (version 12.0.20491.10.21239^57,58^) from Biognosys with default settings applied and were searched against the human Uniprot FASTA database.

### Mass spectrometry bioinformatic and statistical analyses

Mass spectrometry raw files were processed using the MaxQuant software^59^ *(version 1*.*5*.*3*.*34)*. As previously described^47^, peak lists were searched against the human Uniprot FASTA database (*November 2016*), and a common contaminants database (247 entries) by the Andromeda search engine^60^. Pearson correlation analysis, t-test statistics, ANOVA tests, or Fisher’s exact test were performed using the GraphPad Prism 5 software. Protein expression was corrected for age in the following manner. Age was regressed out from the protein expression data using the R function aov(). The residuals from this model were used in subsequent analysis. All other statistical and bioinformatics operations (such as normalization, data integration, annotation enrichment analysis, correlation analysis, hierarchical clustering, principal component analysis, and multiple-hypothesis testing corrections), were run with the Perseus software package (version 1.5.3.0 and 1.6.1.1.) (Tyanova et al, 2016).

## Acknowledgements

We thank all the patients and their families for supporting the progress of science. We gratefully acknowledge the provision of human biomaterial and clinical data from the CPC-M bioArchive and its partners at the Asklepios Biobank Gauting, the Klinikum der Universität München and the Ludwig-Maximilians-Universität München. We thank Igor Paron and Korbinian Mayr for expert support of the proteomics pipeline. We thank Sandy Lösecke and Elisabeth Graf for technical assistance in next generation sequencing and Thomas Schwarzmayr, Thomas Walzthöni and Matthias Heinig for support with High-seq 4000 sequencing raw data and the single cell processing pipeline. We thank Igor Igor Kukhtevich and Robert Schneider for their help with microfluidic devices for dropseq. This work was supported by the German Center for Lung Research (DZL), the Helmholtz Association, the Max Planck Society, and the German Federal Ministry of Education and Research (BMBF), project Single Cell Genomics Network Germany.

## Author contributions

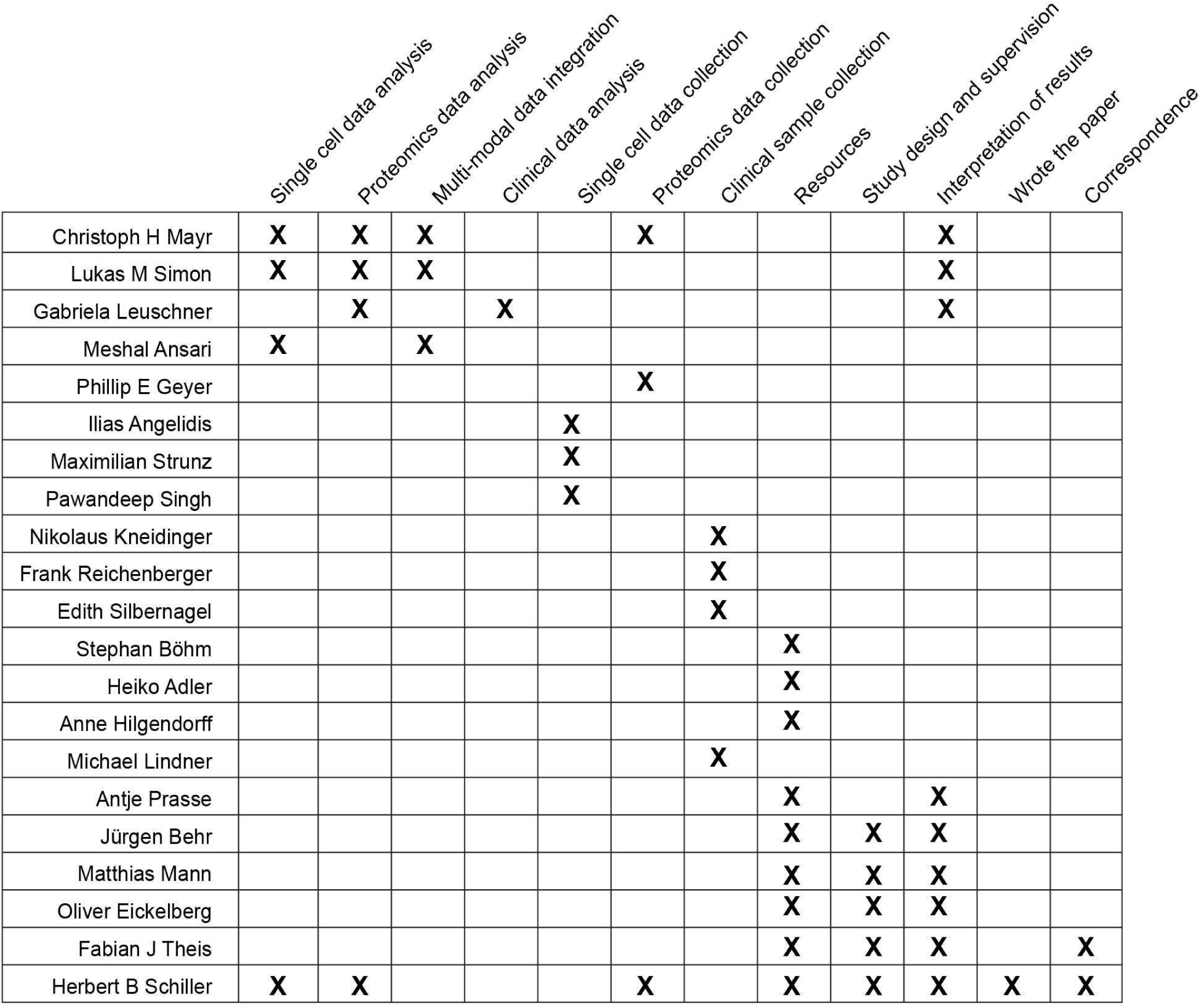

